# Impact of light logger and dosimeter placement on wearability and appeal in real-life settings

**DOI:** 10.1101/2025.06.13.25329548

**Authors:** Johannes Zauner, Anna M. Biller, Manuel Spitschan

## Abstract

Light exposure plays a crucial role in human health, influencing sleep, circadian rhythms, and visual development. As wearable light loggers are increasingly used to monitor real-world light exposure, their design must balance metrological accuracy with comfort and usability. We assessed the perceived wearability and appeal of eight light logger designs and placements (chest pin, wrist, necklace, sleeve collar, collar pin, neck loop, hat pin, and glasses) using an online survey completed by 145 participants (mean age 32 years, 52% female) from the UK, USA, and other countries worldwide. Each design was rated on overall attractiveness, likelihood of wear at home, at work, in public settings, in social contexts, during exercise, expected wear duration, and perceived interference with daily activities. The chest pin received the highest overall ratings, followed by wrist and necklace, while glasses and hat pin placements were rated lowest across most contexts. Likelihood of wear was highest in the home setting and significantly lower in social (β = –0.92 ± 0.08) and work (β = –0.84 ± 0.08) contexts. Compared to the chest pin, other placements were rated up to 2.6 points lower (e.g., glasses: β = –2.62 ± 0.10). Wearing position significantly influenced all ratings (p < 0.001), while sample location and gender showed minimal effects. The thematic analysis of free-text responses revealed concerns around comfort, appearance, stability, and interference with daily activities. These findings highlight the importance of user-centred design and offer practical guidance for developing and deploying wearable light loggers that are acceptable in everyday contexts.

## Background

Exposure to light affects human health and well-being in several ways (Burns et al., 2021; Windred et al., 2024; Zielinska-Dabkowska et al., 2023). In recent decades, growing evidence has linked light exposure—both daytime and nighttime—to circadian and sleep physiology, leading to recommendations for optimal light exposure levels (Brown et al., 2022). Additionally, light exposure and more broadly, the visual diet, have been linked to ocular health, specifically myopia (Marcos, 2025). The neurobiological pathways underlying these so-called ‘non-visual’ effects of light – in contrast to the impact of light on the visual system – have been elucidated, involving a particular set of cells in the retina (Lucas et al., 2014). The intrinsically photosensitive retinal ganglion cells (ipRGCs) express the photopigment melanopsin and signal information about environmental illumination independent of the rods and cones. Consequently, they also signal the time of day to the brain via the retinohypothalamic (RHT) tract. The RHT connects the eye to the suprachiasmatic nucleus (SCN), the neural structure underlying circadian and sleep control.

While most evidence on the non-visual effects of light in humans comes from laboratory studies with a high degree of experimental control, and well-constrained and well-parametrised light, light exposure patterns under natural, everyday conditions are complex, not least due to the behavioural control over light exposure conditions, e.g., stepping out to daylight, or turning on indoor artificial lighting (Biller et al., 2024). Field studies and the translational application of findings on the non-visual effects of light, therefore, pose a particular challenge in the measurement and characterisation of real-world light exposure. Due to this demand, wearable light loggers, sometimes also called dosimeters^1^, have been developed that capture the light exposure of an individual over time (Danilenko et al., 2022; Hartmeyer et al., 2022; Spitschan et al., 2022).

Wearable light loggers are available in a range of form factors, with varying functionalities. The most common type of light logger comes in the form of a wrist-worn actimeter, an accelerometer that tracks rest-activity cycles and, for some models, also light exposure. As the relevant metrological quantity underlying the non-visual effects of light is the activation of the ipRGCs in the retina (CIE, 2018; Lucas et al., 2014; Price & Blattner, 2022; Spitschan, 2019), light exposure should be measured at least in the plane of the eye, which some devices approach by mounting on spectacle frames (Bierman et al., 2005; Hubalek et al., 2016; Stampfli et al., 2023; Wen et al., 2021), with varying acceptability (Balajadia et al., 2023). Importantly, however, the retinal irradiance, rather than the corneal irradiance, is the biological quantity of interest, which is practically impossible to measure due to variations in face shape (Sliney, 1995, 2002), lid opening (Sliney, 1997) and pupil size (Gaddy et al., 1993; Harley & Sliney, 2018; Higuchi et al., 2008; Lazar et al., 2024), changing the effective visual field (Zauner et al., 2023). While light exposure collected on different body locations is correlated (Aarts et al., 2017; Figueiro et al., 2013; Okudaira et al., 1983; Thieden et al., 2000; Wen et al., 2023; Yoshimura et al., 2020), the exact relationships depend on the spatial geometry of an individual’s instantaneous ambient environment and individual behaviour (e.g., covering of the device with sleeves) (Peeters et al., 2020). At the same time, some wearable light loggers may be more usable and, therefore, more tolerable to participants and users under real-world conditions.

As wearable light loggers become central tools in circadian, sleep, and myopia research, understanding user-centred design preferences is key for ensuring high compliance in real-world deployments. To understand the trade-off between the metrological fidelity of light exposure measurements and usability and acceptability in the field, we examined the attractiveness and acceptability for eight distinct light logger designs and placements (chest pin, wrist, necklace, sleeve collar, collar pin, neck loop, hat pin, glasses) in a range of contexts (at home, at work, in a public setting, in social contexts, during exercise) in an online survey to guide the development of an optimal form factor of a wearable light logger.

## Methods

### Development of light logger illustrations

Informed by expert domain knowledge, we developed eight possible designs for wearable light loggers: chest pin, wrist, necklace, sleeve collar, collar pin, neck loop, hat pin, glasses. Some of these designs (chest pin, wrist, necklace, collar pin, glasses) correspond to existing designs of wearable light loggers, while others were conceived specifically for this study, based on broad knowledge of wearable devices in general. These designs were then turned into 2D illustrations in which a male and female mannequin were shown to wear the designs (see **Figure 1, top row**).

**Figure 1.**
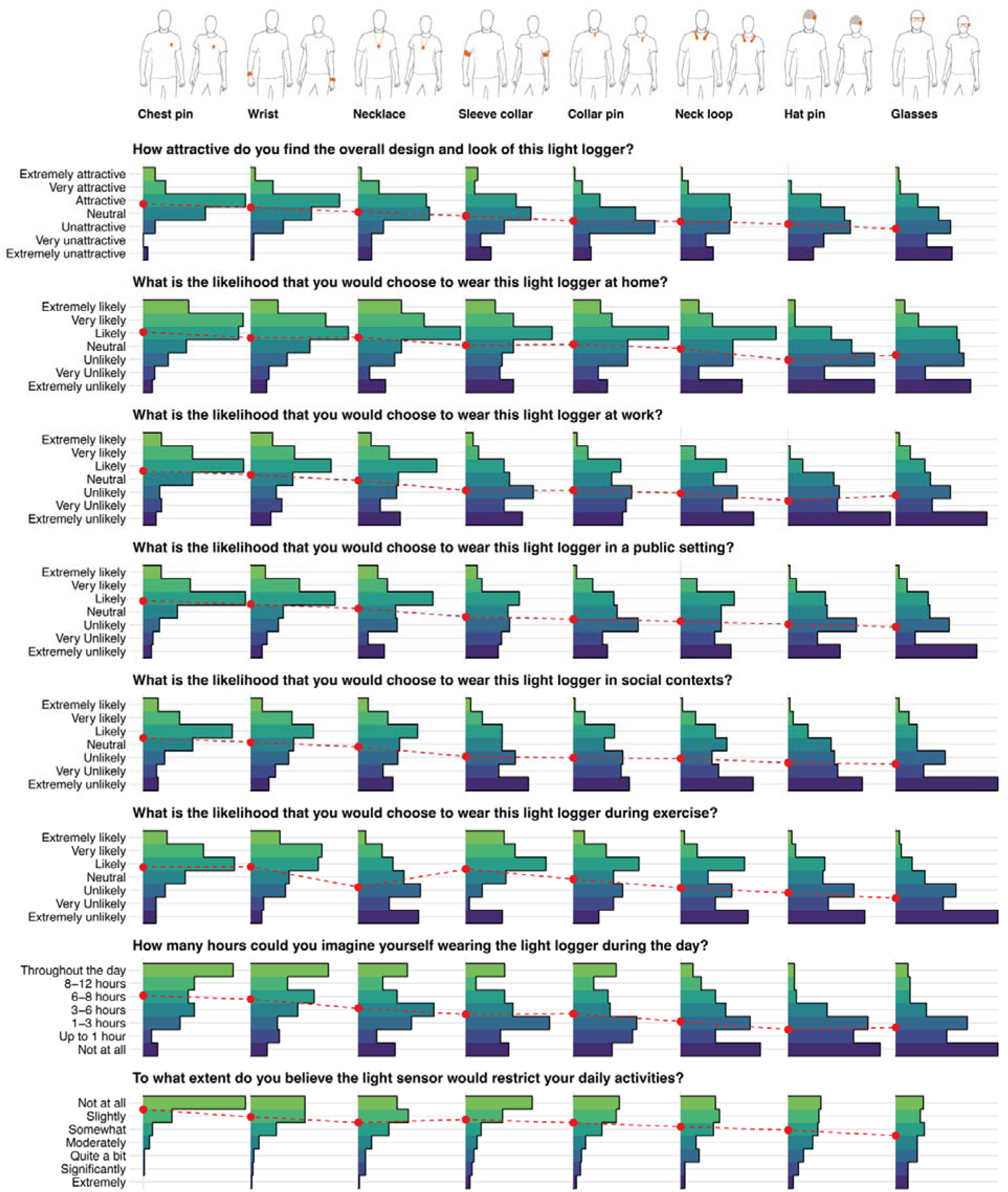
Ratings of light logger wear location. Questions included overall attractiveness and look, different contexts (at home, at work, in a public setting, in social contexts, during exercise), wear duration during the day and interference with daily activities.

### Survey design

#### Recruitment and procedure

We recruited participants through the online platform Prolific (https://www.prolific.com/). Participants were directed to the survey implemented on REDCap (Harris et al., 2019; Harris et al., 2009) (server hosted by the Technical University of Munich), where they also gave their informed consent. Recruitment was completed in three independent waves, recruiting first a US sample, then a UK sample, and then a worldwide sample. The temporal order was determined by platform availability, not study design. Participants were compensated at a rate of £3 for an estimated 20 minutes of their time. For each site, a target sample of n=50 was selected due to resource limitations. All data collection took place in January 2024.

#### Survey questions

We presented the eight light logger designs sequentially and in the same order to participants and asked the following rating questions on a 7-item Likert scale: Question 1: How attractive do you find the overall design and look of this light logger?

`o` Extremely unattractive
`o` Very unattractive
`o` Unattractive
`o` Neutral
`o` Attractive
`o` Very attractive
`o` Extremely attractive

Questions 2-6: What is the likelihood that you would choose to wear this light logger [at home, at work, in a public setting, in social contexts, during exercise]?

`o` Extremely unlikely
`o` Very unlikely
`o` Unlikely
`o` Neutral
`o` Likely
`o` Very likely
`o` Extremely likely

Question 7: How many hours could you imagine yourself wearing the light logger during the day?

`o` Not at all
`o` Up to 1 hour
`o` 1-3 hours
`o` 3-6 hours
`o` 6-8 hours
`o` 8-12 hours
`o` Throughout the day

Question 8: To what extent do you believe the light sensor would restrict your daily activities?

`o` Extremely
`o` Significantly
`o` Quite a bit
`o` Moderately
`o` Somewhat
`o` Slightly
`o` Not at all

For each design, we also asked the participants the following free-form question: “In what situations or activities do you think wearing the light logger might be difficult?”.

#### Demographics data

We asked participants for the following demographic information: age, sex, gender, native speaker status and country of residence and time zone. The entire survey is available as Supplementary Material as a PDF printout and as a REDCap data dictionary.

#### Quality control measures

For each design, we asked participants to describe the light logger in their own words. Additionally, we added three attention-check questions to avoid the automated completion of questionnaires by bots. We removed all data that failed at least one attention check question. The attention check questions were:

- What is 4+5?
- Please type in “nineteen” as a number.
- Please tick the answer “Often”.

### Analytic strategy

#### Quantitative data

Quantitative data were analysed descriptively, inferentially, and visualised using *R* software (R Core Team, 2017). Inferential analysis was performed using cumulative link mixed models with the *ordinal* package in R (Christensen, 2019). The dependent variable was the participant’s rating. Independent variables included wearing position, sex, the sample (location), and context. Context is a derived variable from survey questions two through six, where all data about the wear-likelihood in different situations is pooled, and the context of each question is stored as a variable. This allows exploration of the wearing position in interaction with the context. Participants’ answers for sex and gender were identical for all participants included in inferential analyses containing those covariates. One participant identified as “Other” for sex and gender and was thus excluded from these analyses, as parameters cannot be estimated for a subgroup containing a single sample. Participants were included as random effect.

P-values were obtained by likelihood-ratio tests of the full model with the effect in question against the model without the effect in a stepwise elimination of parameters down to the *null model* with only the random effect. P-values less than or equal to 0.05 were considered significant, with a prior *False-Discovery-Rate* correction (Benjamini & Hochberg, 1995). Beta coefficients provide the parameter estimates with the standard error on the logit scale of the model, compared to the reference level, which is the *Chest pin* position and *home* context.

Two explorative analyses were performed:

1. How does the wearing position influence each question’s rating, with gender and sample location as control variables? As gender might influence how a given wearing position is rated, the full model includes this interaction effect. Note that sex and gender are identical in our sample. The full model in Wilkinson notation is thus:

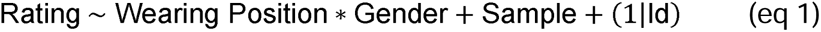

Post-hoc analyses were performed to identify significantly different wearing positions, also with a *False-Discovery-Rate* correction.

2. How does the context of wear (work, social, etc.) influence the ratings, dependent on the wearing position? The full model in Wilkinson notation is thus:

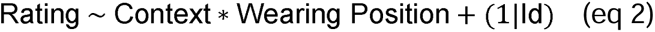

Gender was not included in this second part for reasons of computational stability, speed, and redundancy with the first explorative analysis, where gender differences are explored within each context.

All scripts used for descriptive and inferential analysis are available as Quarto HTML files in the supporting information.

#### Qualitative data

Qualitative responses to survey questions were analysed using the large-language model (LLM) Gemini 2.5 Pro Preview 05-06 (Google, Mountain View, USA). The analysis was performed in a three-step process. In the first step, qualitative answers were extracted from the survey and brought into a long format of one answer per question and design per row. In the second step, performed by the LLM, themes were identified, and each participant’s answer in the 16 design-specific questions (8 designs with 2 questions each) and two general questions was assigned to a theme (or sorted to a “non-theme” bin). If multiple themes were touched by a single answer, these answers could be assigned multiple times. These data were exported from the LLM for the final step in R software (see Quarto HTML in the supporting information). In this step, it was checked whether each of the theme-quotes matches a participant’s answer and was not produced through LLM hallucination. This was the case in all quotes. Further, it was checked whether all answers from the survey were part of the thematic analysis, i.e., no answers were left out. This was the case. Finally, multiple rounds of random samples of quotes were manually checked against the themes to check their validity and appropriateness within the theme. Summaries within and across themes were also supported by the LLM and checked for validity against the dataset.

## Results

### Participant demographics

We were able to recruit a total of 162 participants. One participant did not give their consent and was removed. 16 participants did not pass the attention checks and were removed. One participant did not complete the survey and was removed. The remaining 145 respondents originate from the survey samples as follows: n=47 (UK), n=49 (USA), and n=49 (worldwide). The demographic results for the total sample and the sub-samples are given in **Table 1**. In brief, the average age of respondents was 32 years (range 18-81); 75 (52%) were female, 69 were male (48%), and one person identified as ’Other’ (0.7%). The majority of our participants (95, 66%) were native English speakers, with a larger proportion in the UK and USA samples (85% [UK] and 86% [USA], respectively). In terms of occupation, most participants reported being in full-time employment (46%), followed by people working part-time (19%) or being in education (18%). Our global sample shows a diverse distribution of 17 countries of origin (**Table S1**).

**Table 1.** Sample demographics. Answers for gender are identical to sex.

| Demographics | Overall, N = 145<sup>1</sup> | Sample locations |  |  |
| --- | --- | --- | --- | --- |
|  |  | UK, N = 47<sup>1</sup> | USA, N = 49<sup>1</sup> | Worldwide, N = 49<sup>1</sup> |
| Age (yrs) | 32 (18-81) | 33 (18-75) | 39 (19-72) | 27 (18-81) |
| Sex |  |  |  |  |
| Female | 75 (52%) | 26 (55%) | 25 (51%) | 24 (49%) |
| Male | 69 (48%) | 21 (45%) | 24 (49%) | 24 (49%) |
| Other | 1 (0.7%) | 0 (0%) | 0 (0%) | 1 (2.0%) |
| Native English speaker | 95 (66%) | 40 (85%) | 42 (86%) | 13 (27%) |
| Occupation |  |  |  |  |
| In education | 26 (18%) | 6 (13%) | 7 (14%) | 13 (27%) |
| Part-time working | 28 (19%) | 6 (13%) | 14 (29%) | 8 (16%) |
| Full-time working | 67 (46%) | 26 (55%) | 19 (39%) | 22 (45%) |
| Unemployed | 17 (12%) | 8 (17%) | 7 (14%) | 2 (4.1%) |
| Rest | 7 (4.8%) | 1 (2.1%) | 2 (4.1%) | 4 (8.2%) |
| <sup>1</sup> Median (Minimum-Maximum); n (%) |  |  |  |  |

### Attractiveness, usability and acceptability ratings across designs, contexts and samples

#### Descriptive analysis

The results of this study are shown in **Figure 1** and tabulated in **Table S2**. It is important to note that questions two through six ask about the likelihood of a participant choosing a given design. Therefore, the term likelihood in the results section refers to the phrasing of the question, not the stricter statistical definition.

Overall, the *Chest pin* position was rated most favorable, followed by *Wrist*, *Necklace*, *Sleeve collar*, *Collar pin*, *Neck loop*, *Hat pin*, and, least favorable, *Glasses*. The order from best to lowest ranking is:

1. Chest pin
2. Wrist
3. Necklace
4. Sleeve collar
5. Colar pin
6. Neck loop
7. Hat pin
8. Glasses

This order holds across all questions, with very few exceptions. Notably, people rated the *Necklace* position less favorable for exercise than the *Sleeve collar* or *Collar pin,* and rated the *Necklace* as slightly more restricting than the *Sleeve collar* during daily activities. Further, the *Hat pin* position was rated lower than *Glasses* for wear at home and work.

#### Inferential analysis

The first branch of the exploratory analysis examined how the wearing position influences each question’s rating, with sex/gender and sample location as control variables. The wearing position contributes significantly to explain ratings in all questions (all p < 0.001), with the basic order being the same as described in the descriptive analysis results above. **Figure 2** shows which wearing positions differ significantly from one another in their rating. Sample location had no effect (all p ≥ 0.077; **Figure S1**), and neither did gender (all p ≥ 0.61). The interaction of gender and wearing position was significant in the questions about the social context (p = 0.032) and for the wearing duration (p = 0.005). In both cases, the interaction significantly affected only a few wearing positions. Namely, in a social context, male participants rated the Sleeve collar position about one step better than female participants (*β* = 1.2 ± 0.43), leading to about the same rating as the *Necklace* position. For wear duration, men would be willing to wear the *Neck loop* (*β* = 1.33 ± 0.44) or the *Hat pin* (*β* = 1.51 ± 0.44) about one to one and a half steps longer than females. **Figure 2** shows where the interaction of sex and wearing position leads to further significant differences in rating compared to the wearing position alone.

**Figure 2.**
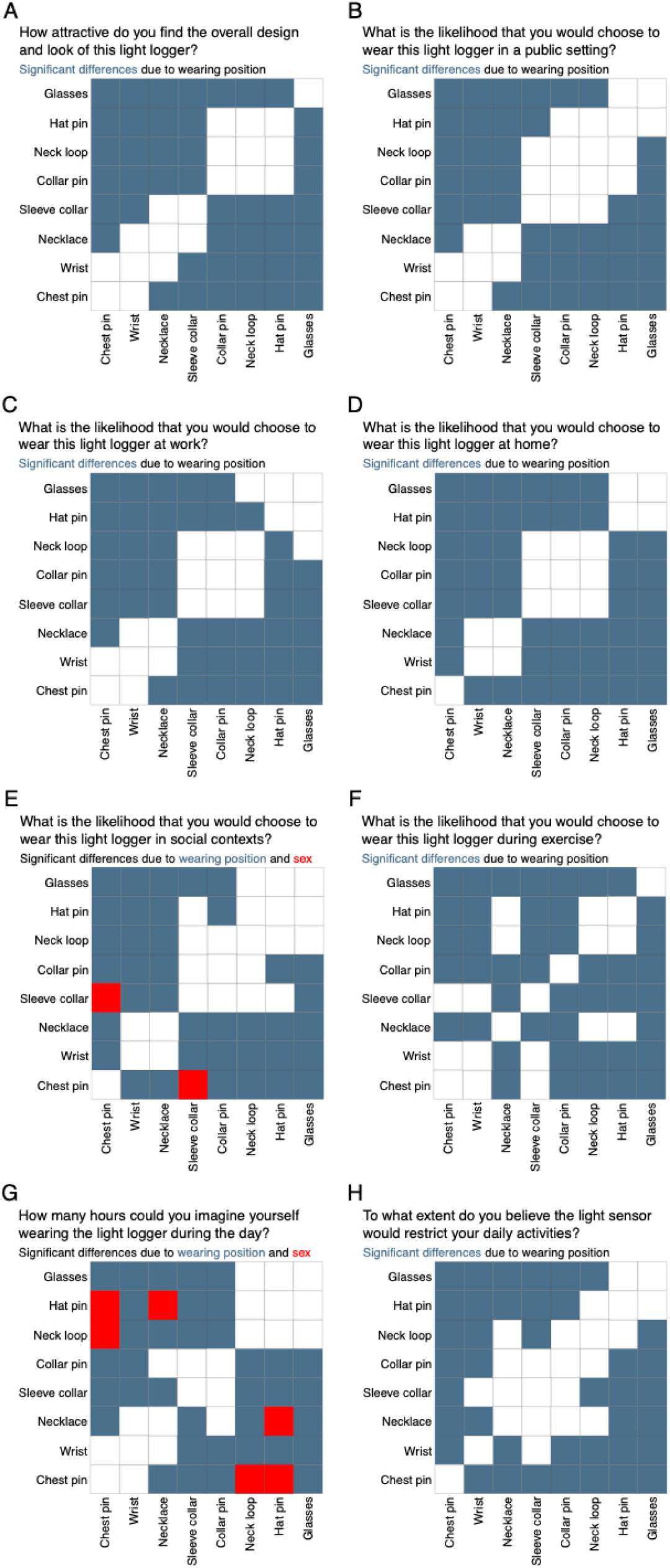
Statistically significant differences in ratings between wearing position pairs. Blue squares indicate that ratings differ significantly between these pairs, and red squares further indicate a significant difference in rating depending on sex/gender for these pairs. Note that participants’ answers for sex and gender were identical in our sample.

The second branch of the exploratory analysis examined how context influences the ratings. This analysis thus excluded the questions on attractiveness, wearing duration, and restriction, and only included the questions about the likelihood of choosing to wear a particular design in a public setting, at work, at home, in social contexts, or during exercise. In this model, context, wearing position, and their interaction were significant (all p < 0.001). Looking at the singular effect of context, people are more willing to rate any design higher (more favorable) at home, compared to a public (*β* = −0.45 ± 0.07), exercise (*β* = −0.45 ± 0.08), work (*β* = −0.84 ± 0.08), or social setting (*β* = −0.92 ± 0.08), where a *β* of about −1 equals to one rating level lower (less favorable). **Table 2** shows the likelihood of wear ratings depending on context. Looking at the singular effect of wearing position, people are more willing to rate the *Chest pin* position higher (more favorable) regardless of context, compared to *Wrist* (*β* = −0.34 ± 0.09), *Necklace* (*β* = −1.02 ± 0.09), *Sleeve collar* (*β* = −1.40 ± 0.10), *Collar pin* (*β* = −1.62 ± 0.10), *Neck loop* (*β* = −1.99 ± 0.10), *Hat pin* (*β* = −2.60 ± 0.10), or *Glasses* position (*β* = −2.62 ± 0.10), where a *β* of about –1 equals one rating level lower (less favourable). **Table 3** shows the likelihood of wear ratings depending on wearing position. Looking at the combined effect of context and wearing position, **Table 4** shows the predicted mode rating of each wearing position depending on context. The table summarises concisely that the *Chest pin* and *Wrist* are generally accepted wearing positions (mode: *likely*). The *Necklace* is mostly accepted but rated *unlikely* for exercise. *Sleeve collar*, *Collar pin*, and *Neck loop* are accepted for the home context and are partly preferable for exercise but are rated *unlikely* for other contexts. The *Hat pin* and *Glasses* are the lowest rated wearing positions overall and are rated *extremely unlikely* to be worn in a work or social setting.

**Table 2.** Model prediction for the likelihood of wear ratings depending on context. The colouring emphasises the range of numeric values.

| Likelihood of wear ratings depending on context |  |  |  |  |  |  |  |
| --- | --- | --- | --- | --- | --- | --- | --- |
| Parameters | Extremely unlikely | Very Unlikely | Unlikely | Neutral | Likely | Very likely | Extremely likely |
| home | 8% | 9% | 17% | 19% | 30% | 13% | 4% |
| public | 13% | 12% | 20% | 19% | 25% | 9% | 3% |
| exercise | 13% | 12% | 20% | 19% | 25% | 9% | 3% |
| work | 18% | 14% | 22% | 18% | 20% | 6% | 2% |
| social | 19% | 15% | 23% | 17% | 19% | 6% | 2% |

**Table 3.** Model prediction for the likelihood of wear ratings depending on wearing position. The colouring emphasises the range of numeric values.

| Likelihood of wear ratings depending on wearing position |  |  |  |  |  |  |  |
| --- | --- | --- | --- | --- | --- | --- | --- |
| Parameters | Extremely unlikely | Very Unlikely | Unlikely | Neutral | Likely | Very likely | Extremely likely |
| Chest pin | 3% | 4% | 10% | 16% | 39% | 21% | 7% |
| Wrist | 4% | 5% | 13% | 19% | 37% | 17% | 5% |
| Necklace | 8% | 9% | 19% | 22% | 30% | 10% | 3% |
| Sleeve collar | 11% | 12% | 23% | 22% | 25% | 7% | 2% |
| Collar pin | 13% | 13% | 24% | 21% | 21% | 6% | 1% |
| Neck loop | 18% | 16% | 25% | 19% | 17% | 4% | 1% |
| Hat pin | 28% | 20% | 24% | 14% | 10% | 2% | 1% |
| Glasses | 29% | 21% | 24% | 14% | 10% | 2% | 1% |

**Table 4.**
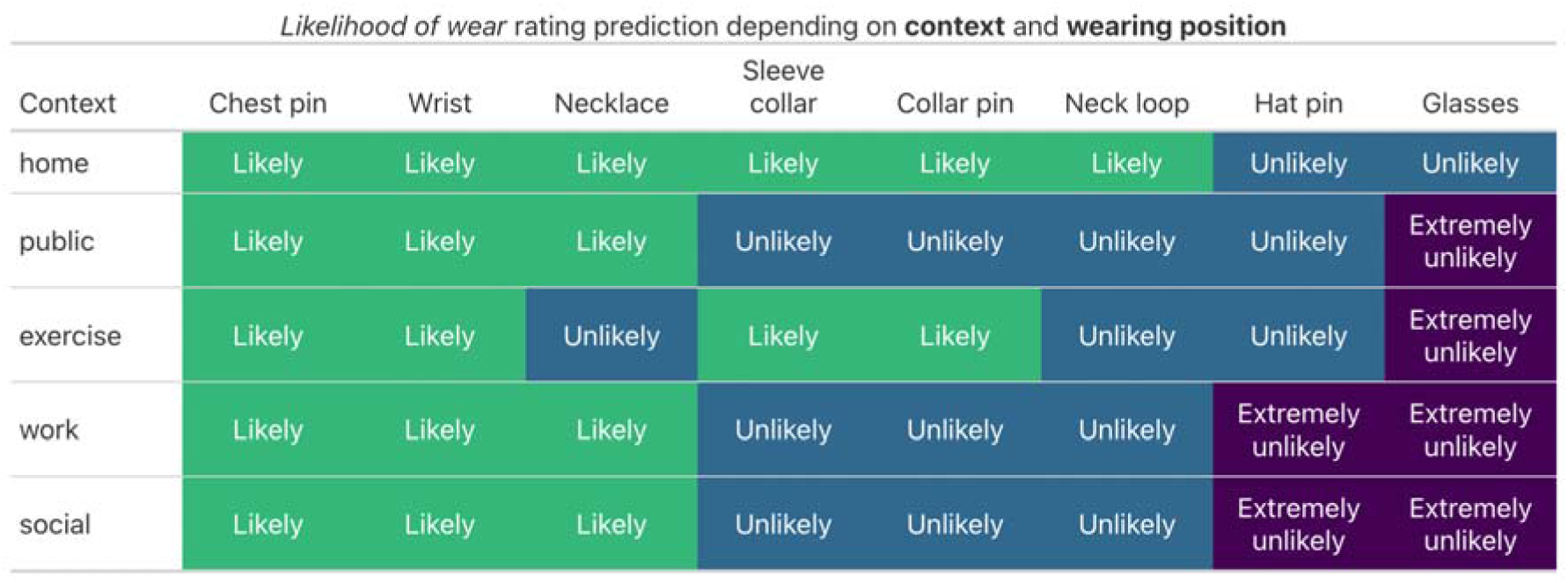
Model prediction for the mode rating depending on context and wearing position. The colouring emphasises the range of the rating.

#### Qualitative results

The analysis identified three overarching themes and several specific sub-themes influencing the wearability and acceptability of the light logger designs. All themes and the number of instances these were mentioned by users can be found in Table 5.

**Table 5.**
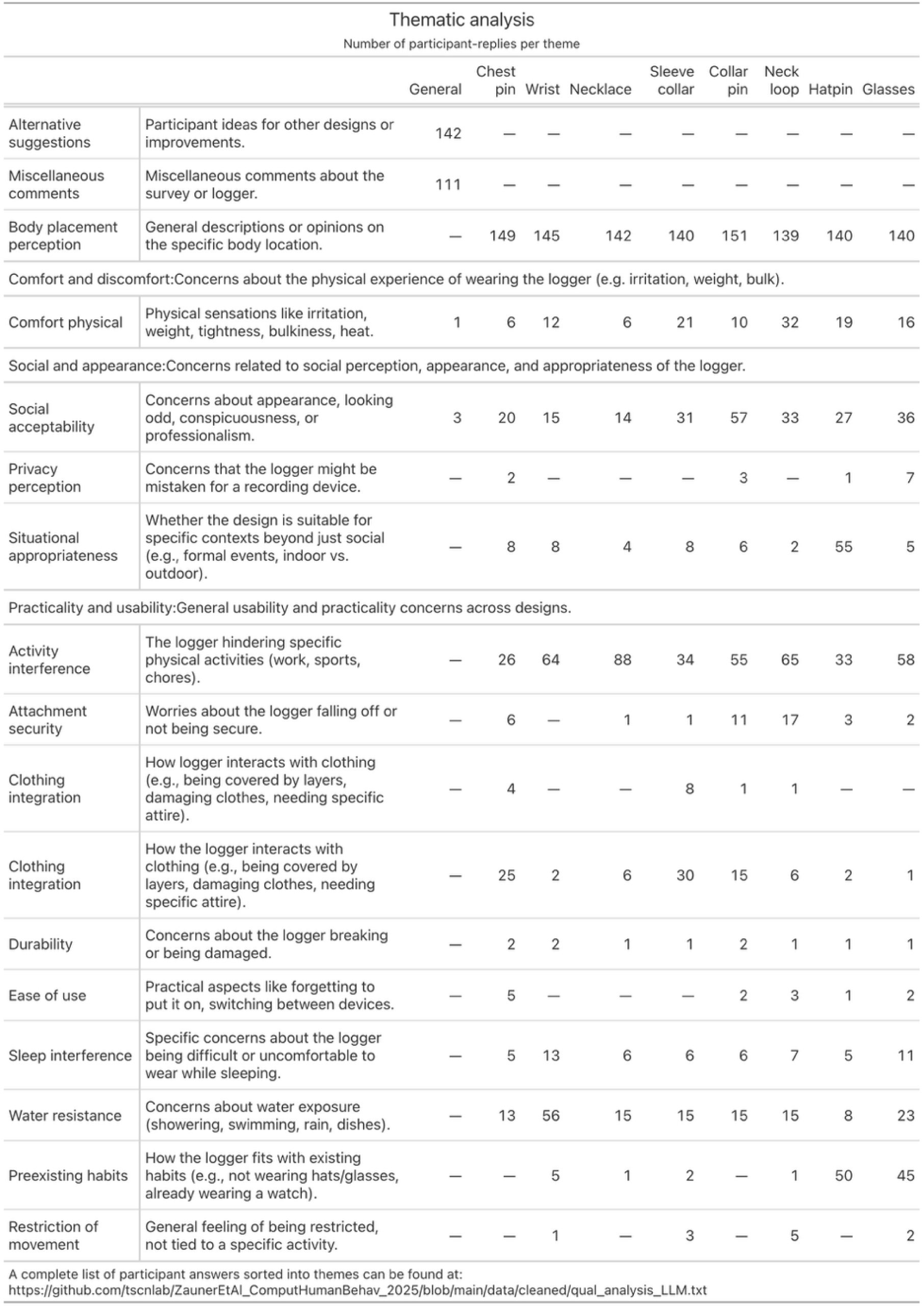
Thematic analysis.

### Practicality & usability

This was a dominant theme, reflecting concerns about how well the logger would function in daily life.

- **Activity interference:** A major concern across almost all designs. Participants frequently cited difficulties wearing loggers during physical work (e.g., “I work for a moving company”), exercise (“intense exercise”, “rock climbing”, “running”, “lifting weights”), or daily chores (“cooking”, “doing dishes”).

- *Wrist:* “my main hobby is rock climbing”, “typing or using a computer”.
- *Collar-pin, Chest-pin, Necklace:* Concerns about them falling off or hindering movement during “intense exercise”.
- *Glasses:* “working out”, “anything with a lot of movement”.
- *Neck-loop:* “Anything with a lot of movement”, “Doing exercises”.
- **Sleep interference:** A frequently cited concern for nearly all logger designs was the anticipated difficulty or discomfort of wearing them during sleep. Participants explicitly mentioned “Sleep hours” as problematic for designs like the *collar-pin, glasses, hat-pin, neck-loop, chest-pin, necklace,* and *sleeve-collar*. The *wrist*logger also prompted comments such as, “light logger might be difficult wearing when sleeping. would not be able to sleep on my hand properly”, and general difficulty “when sleeping or laying in bed”. For the *glasses* design, one participant stated, “I would not wear it to sleep”. The *hat-pin* was also singled out: “Exercising, sleeping, taking a shower, swimming will be difficult to do”. This indicates that achieving continuous wear through the night presents a significant design challenge across most form factors, as users prioritize comfort and unhindered rest.
- **Attachment security:** Primarily for pin/clip-based designs (collar-pin, chest-pin) and loose-fitting designs (neck-loop, necklace). Participants worried about devices “falling off easily” or not being “stable”.
- **Clothing integration:** How the logger interacts with clothing was a common point.

- *Wrist, Sleeve-collar:* Obstruction by “long sleeved shirt or jacket”, “cold climates where you wear lots of layers”,
- *Chest-pin, Collar-pin:* Being covered by a “coat”, or issues with “different shaped clothing”. Some worried about pins “damaging clothing”.
- **Water resistance:** Frequently mentioned for designs in direct skin contact or likely to get wet. “Showering/bathing”, “swimming”, “rainy weather”, and “doing dishes” were common examples where participants would be concerned or remove the device. This applied strongly to *wrist, glasses, hat-pin, neck-loop, chest-pin, necklace, sleeve-collar*.
- **Durability:** Some participants, especially for the *wrist* design, drew parallels to other wearables, expressing concerns like, “I used to wear an apple watch that I destroyed pretty quickly”.
- **Ease of use & pre-existing habits:**
- *Glasses:* “if you do not already wear glasses it forces you to”, “having to switch out” with regular glasses.
- *Hat-pin:* “I rarely wear hats”, “any work setting or anywhere it would be uncomfortable / unfit to wear a beanie”.
- *Wrist:* For some, if it’s “not also a watch, I would likely find it uncomfortable to wear a watch like device on both arms”.
- Forgetting to transfer pin/clip devices when changing clothes was also a minor ease-of-use concern.

### Social & appearance concerns

The aesthetic and social perception of the loggers significantly influenced acceptability.

- **Social acceptability:** Many designs were deemed “conspicuous”, “silly”, “ugly”, “awkward”, or “unfashionable”, particularly for “social settings”, “formal events”, or “work”.

- *Collar-pin:* “looks like a bad/conspicuous spot so wearing it socially wouldn’t be ideal.”
- *Glasses:* “attention drawn to my face”.
- *Hat-pin:* “unfashionable in work and social situations”.
- *Neck-loop:* “Ugly”, “silly in my opinion”.
- *Sleeve-collar:* “looks like exercise or medical equipment”.
- **Privacy Perception:** Some designs, notably *glasses*, *chest-pin*, and *collar-pin*, raised concerns about being mistaken for a “hidden camera or recording devices”, making others uncomfortable.
- **Situational appropriateness:** Beyond general social settings, some designs were seen as inappropriate for specific contexts.

- *Hat-pin:* “indoors - where hats are uncommon”, “not polite”.
- *Chest-pin/Collar-pin:* Concerns about looking “unprofessional” or drawing unwanted questions.

### Physical comfort & discomfort

- **Comfort/physical sensation:** Issues included potential “skin irritation”, “extra weight” (especially for *glasses*), feeling “clunky”, “bulky”, “heavy”, or generally “uncomfortable”.

- *Wrist:* “irritated by texture/weight on your wrist”.
- *Collar-pin:* “Seems uncomfortable”.
- *Neck-loop:* “irritating to have some weight / thing hang around your neck”.
- *Necklace:* “annoying […] something rubbing against your neck”.
- *Sleeve-collar:* “irritating on my upper arm”.
- *Hat-pin:* Hat itself becoming “hot” or “messing up hair”.

### Design-specific perceptions & alternative suggestions

- **Wrist:** Often seen as the most familiar (“like a watch”, “Fitbit type”), but concerns about bulkiness, water, and interference with tasks/other watches existed.
- **Collar-pin & Chest-pin:** Liked by some for potential discreetness if small, but others found the placement odd, worried about security, clothing damage, or looking like a microphone. The *chest-pin* design can be inconspicuous (“Doesn’t appear as though it would interfere”).
- **Glasses:** Concerns included weight, bulk, peripheral vision, and social awkwardness. A potential issue are wearers who don’t normally wear glasses.
- **Hat-pin:** Limited by the necessity of wearing a hat, making it unsuitable for many situations (indoors, warm weather, formal settings).
- **Neck-loop:** Frequently criticized as “ugly”, “silly”, “cumbersome”, and prone to falling off.
- **Necklace:** Seen as potentially acceptable if discreet, but concerns about swinging during activity and tangling were common.
- **Sleeve-collar:** Mixed reactions; some saw it as non-intrusive, others as restrictive, uncomfortable, or socially awkward (“weird arm band”).
- **Alternative Suggestions:** A strong recurring suggestion was for **smaller, more discreet designs**. Specifically, a **ring** was mentioned by some participants (“maybe a ring because a lot of people already wear rings,” “a ring version […] low-key”). **Earrings** or **hair clips/accessories** were also suggested, aiming for fashion integration. Integration into existing clothing (“fabric integration”, “stickers”) or multi-functional devices (e.g., combined with a smartwatch) were other ideas. Some suggested a “small clip (the size of a stamp) that can be attached anywhere”.

## Discussion

### Differences in attractiveness, usability and acceptability in light logger design and placement

Our study revealed that there are marked differences in the subjective prospective attractiveness, usability and acceptability of wearable light loggers as assessed in a survey based on 2D design prototypes. We found clear differences in these properties across device design and placement and between contexts. The findings suggest that in order to optimise wearable light loggers for field use and translational applications, special care must be taken when selecting a specific wearable design. When comparing ratings, the *chest pin* design was most favoured by participants, followed by *wrist*, *necklace*, and *sleeve collar.* The least favoured was the *glasses* design, followed by *hat pin*, *neckloop*, and *collar pin*. This order was very stable across situations, with only slight deviations connected to exercise (where the loose *necklace* design was rated worse than the *sleeve collar* and *collar pin*) or gender (where the *sleeve collar* position was rated better by male participants in a social context). Notably, even the best-rated design, the *chest pin* raised concerns regarding conspicuity, physical discomfort, and suitability for different clothing types. Looking at qualitative feedback, the findings highlight a clear tension between the need for a light logger to be exposed to ambient light and the user’s desire for discretion, comfort, and seamless integration into daily life. Practicality concerns, such as interference with activities and water resistance, are paramount. Social acceptability and physical comfort are also major determinants of willingness to wear a device continuously. No single design was universally accepted, but those resembling familiar items (wrist-watch) or offering high potential for discretion (a very small chest-pin or a sleek necklace) were generally preferred over obtrusive or habit-changing designs (glasses for non-wearers, hat-pins, neck-loops). The strong preference for miniaturised and integrated alternatives (rings, earrings) suggests that future light logger development should prioritise unobtrusiveness and aesthetic appeal, potentially through co-design with users. Addressing concerns about activity interference, attachment security, and water resistance will also be critical for achieving high wearability and compliance in research settings.

### Use of subjective measures

We probed the attractiveness, usability and acceptability using subjective ratings based on 2D illustrations. This approach allows for probing preferences but does not replace user testing through physical devices. Future research needs to examine the usability of different physical light logger designs, e.g., through non-functional prototypes as a starting point, through qualitative interviews, focus groups, and field testing with both short and longitudinal wear time. Data from our survey-based approach could then be compared and confirmed, or disconfirmed, to the empirical data collected in such an approach. As wearable light loggers continue to be developed and receive attention also in clinical contexts, such studies will become of increasing importance.

### Single-context evaluation

In this study, we asked participants to evaluate the wearable light logger designs and placement locations across a series of contexts, namely at home, at work, in public setting, in social contexts, and during exercise. However, even within these ‘macro-contexts’, there may be significant differences in the appropriateness of different designs and placements. For example, swimming or underwater exercise certainly falls into the category of ‘exercise’, but research-grade wearable light loggers can typically not be used in these contexts. A more fine-grained evaluation of different activities may be necessary to understand differences in different types of exercise or physical activities (e.g., running vs. yoga). Similarly, it may be the case that for light loggers only worn in occupational settings, some specific designs will be more appropriate than others. Season might be a major additional factor, as clothing and clothing-related behaviour changes quite strongly between winter and summer. Additionally, the length during which these designs are worn (not just the hours in the day, which was asked in the survey) could change people’s perception of the designs.

### Population-specific considerations

As our sample included only adults, our findings may not extend to other populations. Children are likely to have different needs and preferences when it comes to wearable devices, influenced by factors like body size, clothing, and daily routines. The same holds for other groups, such as older adults or people in different cultural or occupational contexts. These differences underline the importance of considering the specific needs of each target group when developing wearable devices. Ideally, this should be done in collaboration with the people who will actually use them through co-design and co-creation approaches.

## Conclusion

In this study, we examined subjective usability and acceptability of eight distinct designs and placement locations for wearable light loggers. Our results indicate that not all wearable light loggers are equal, and that there are marked differences in estimated attractiveness, usability and acceptability which also depended on context and sex. We did not see differences based on the location of the participants (UK, USA, worldwide). Future research needs to be done to examine behavioural compliance with different physical light logger designs and placement locations to optimise the trade-off between metrological fidelity and use of wearable light loggers in field and translational contexts.

## Statements

### Ethics approval and consent to participate

This study was reviewed and approved by the TUM Ethics Committee (2024-26-W-CB, 23 January 2024). Participants gave their informed consent to participate.

### Consent for publication

Not applicable.

### Availability of data and materials

All data and materials can be downloaded under a Creative Commons license (CC-BY-NC-ND) from our GitHub repository: https://github.com/tscnlab/ZaunerEtAl_biorXiv_2025

### Competing interests

J.Z. and M.S. participated in the MAX!mize start-up incubator program on a start-up on the development of a wearable light logger and the behavioral modification of light exposure by Max Planck Innovation. M.S. declares the following potential conflicts of interest in the past five years (2021–2025). **Academic roles:** Member of the Board of Directors, *Society of Light, Rhythms, and Circadian Health (SLRCH)*; Chair of *Joint Technical Committee 20 (JTC20)* of the *International Commission on Illumination (CIE)*; Member of the *Daylight Academy*; Chair of *Research Data Alliance Working Group Optical Radiation and Visual Experience Data*. **Remunerated roles:** Speaker of the Steering Committee of the *Daylight Academy*; Ad-hoc reviewer for the *Health and Digital Executive Agency* of the *European Commission*; Ad-hoc reviewer for the *Swedish Research Council*; Associate Editor for *LEUKOS*, journal of the *Illuminating Engineering Society*; Examiner, *University of Manchester*; Examiner, *Flinders University*; Examiner, *University of Southern Norway*. **Funding:** Received research funding and support from the *Max Planck Society*, *Max Planck Foundation*, *Max Planck Innovation*, *Technical University of Munich*, *Wellcome Trust*, *National Research Foundation Singapore*, *European Partnership on Metrology*, *VELUX Foundation*, *Bayerisch-Tschechische Hochschulagentur (BTHA)*, *BayFrance (Bayerisch-Französisches Hochschulzentrum)*, *BayFOR (Bayerische Forschungsallianz)*, and *Reality Labs Research*. **Honoraria for talks:** Received honoraria from the *ISGlobal*, *Research Foundation of the City University of New York* and the *Stadt Ebersberg, Museum Wald und Umwelt*. **Travel reimbursements**: *Daimler und Benz Stiftung*. **Patents:** Named on European Patent Application EP23159999.4A (“*System and method for corneal-plane physiologically-relevant light logging with an application to personalized light interventions related to health and well-being*”). With the exception of the funding source supporting this work, M.S. declares no influence of the disclosed roles or relationships on the work presented herein. The funders had no role in study design, data collection and analysis, decision to publish or preparation of the manuscript.

## Funding

This work was supported by the Max Planck Society in the form of a free-floating Max Planck Research Group, and the MeLiDos Project. The project (22NRM05 MeLiDos) has received funding from the European Partnership on Metrology, co-financed by the European Union’s Horizon Europe Research and Innovation Programme and by the Participating States. All authors are part of the project. The project directly funds the position of J.Z. The funders had no role in study design, data collection and analysis, decision to publish, or preparation of the manuscript.

## Authors’ contributions

Conceptualisation: AB, JZ, MS

Data Curation: MS

Formal Analysis: AB, JZ, MS

Funding Acquisition: MS

Investigation: AB, JZ, MS

Methodology: AB, JZ, MS

Project Administration: MS

Resources: –

Software: JZ

Supervision: –

Validation: –

Visualisation: JZ

Writing – Original Draft Preparation: MS

Writing – Review & Editing: AB, JZ, MS

## Acknowledgments

We thank Max Dobberkau for the illustrations of the light loggers, and Meike Rensen for an early design of the questionnaire.

---

**Figure S1.**
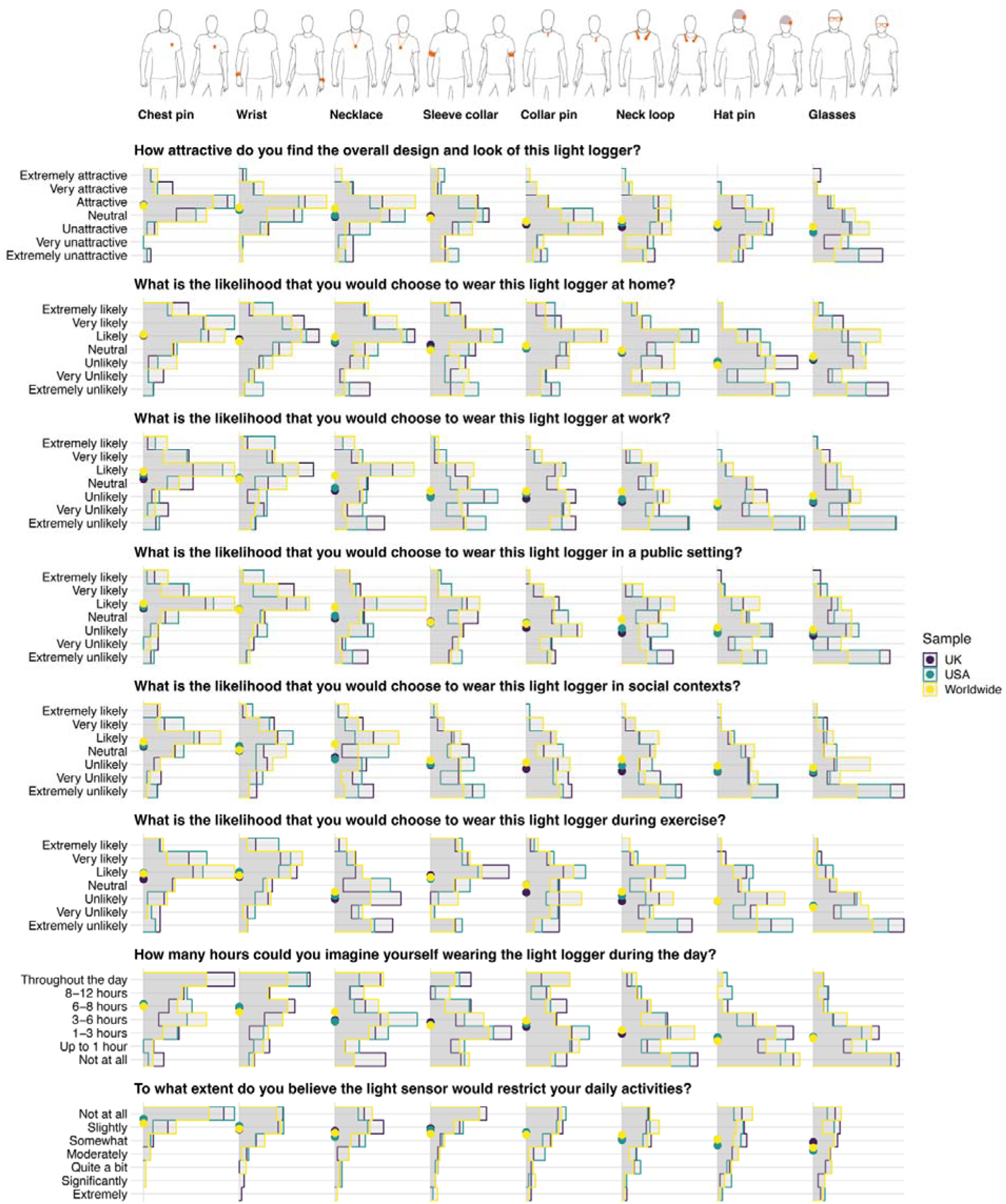
Light logger wear location ratings by sample locations.

**Table S1.** Countries of origin of the sample.

| Countries | N = 145<sup>1</sup> |
| --- | --- |
| United States | 49 (34%) |
| United Kingdom | 46 (32%) |
| Italy | 10 (6.9%) |
| Poland | 9 (6.2%) |
| South Africa | 9 (6.2%) |
| Portugal | 6 (4.1%) |
| Spain | 3 (2.1%) |
| Germany | 2 (1.4%) |
| Greece | 2 (1.4%) |
| Hungary | 2 (1.4%) |
| Australia | 1 (0.7%) |
| Belgium | 1 (0.7%) |
| Canada | 1 (0.7%) |
| Czech Republic | 1 (0.7%) |
| Estonia | 1 (0.7%) |
| Ireland | 1 (0.7%) |
| Sweden | 1 (0.7%) |

<sup>1</sup> n (%) **Table S1. Countries of origin of the sample.**
<sup>1</sup> n (%)

**Table S2.**
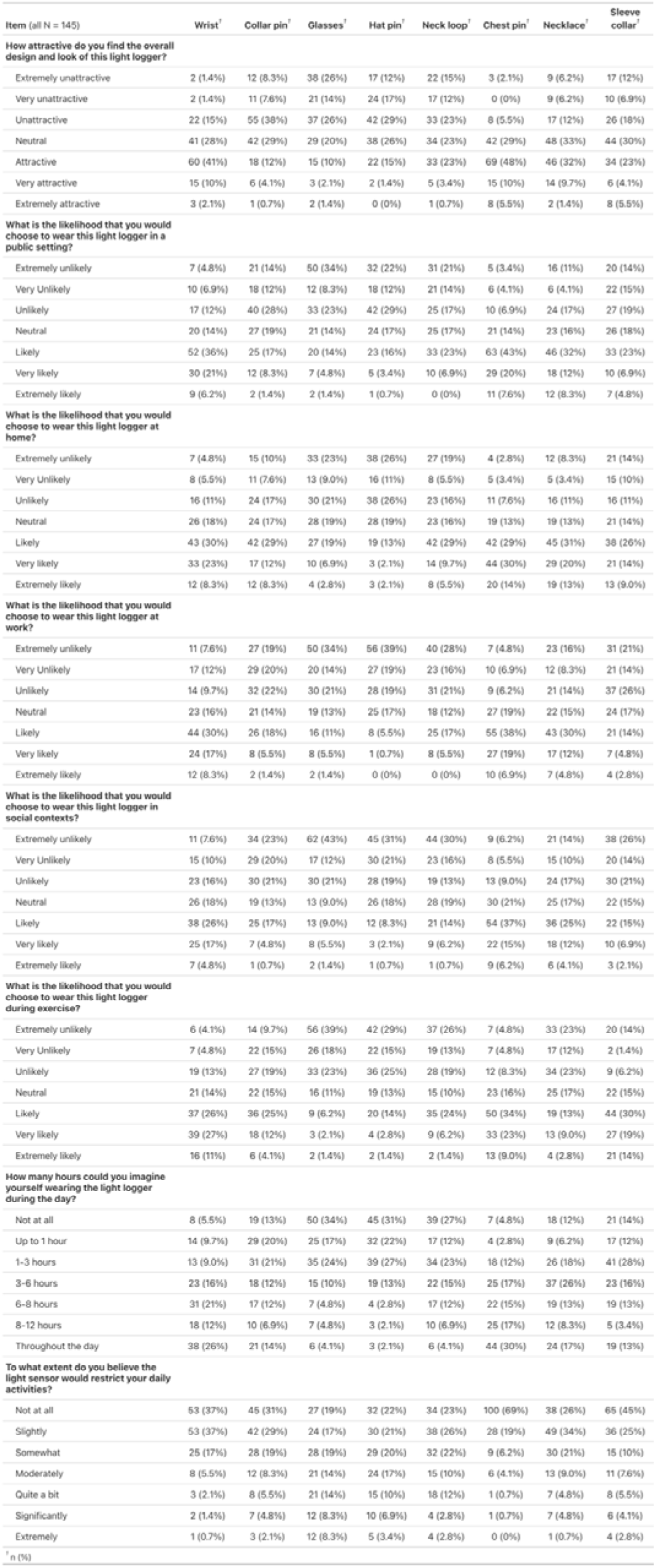
Descriptive statistics of the rating responses.

## Footnotes

1 In the context of this article, we will use the term “wearable light logger” for any electronic devices that measure light exposure in personal fashion. Some manufacturers may use other terms for such devices. The term “dosimeter” is sometimes used for devices or instruments that measure a total dose, i.e. integrated over time.

## References

1. Aarts, M. P. J., van Duijnhoven, J., Aries, M. B. C., & Rosemann, A. L. P. (2017). Performance of personally worn dosimeters to study non-image forming effects of light: Assessment methods. Building and Environment, 117, 60–72. 10.1016/j.buildenv.2017.03.002

2. Balajadia, E., Garcia, S., Stampfli, J., Schrader, B., Guidolin, C., & Spitschan, M. (2023). Usability and Acceptability of a Corneal-Plane alpha-Opic Light Logger in a 24-h Field Trial. Digit Biomark, 7(1), 139–149. 10.1159/000531404

3. Benjamini, Y., & Hochberg, Y. (1995). Controlling the False Discovery Rate: A Practical and Powerful Approach to Multiple Testing. Journal of the Royal Statistical Society: Series B (Methodological), 57(1), 289–300. 10.1111/j.2517-6161.1995.tb02031.x

4. Bierman, A., Klein, T. R., & Rea, M. S. (2005). The Daysimeter: a device for measuring optical radiation as a stimulus for the human circadian system. Measurement Science and Technology, 16(11), 2292–2299. 10.1088/0957-0233/16/11/023

5. Biller, A. M., Balakrishnan, P., & Spitschan, M. (2024). Behavioural determinants of physiologically-relevant light exposure. Commun Psychol, 2(1), 114. 10.1038/s44271-024-00159-5

6. Brown, T. M., Brainard, G. C., Cajochen, C., Czeisler, C. A., Hanifin, J. P., Lockley, S. W., Lucas, R. J., Munch, M., O’Hagan, J. B., Peirson, S. N., Price, L. L. A., Roenneberg, T., Schlangen, L. J. M., Skene, D. J., Spitschan, M., Vetter, C., Zee, P. C., & Wright, K. P., Jr. (2022). Recommendations for daytime, evening, and nighttime indoor light exposure to best support physiology, sleep, and wakefulness in healthy adults. PLoS Biol, 20(3), e3001571. 10.1371/journal.pbio.3001571

7. Burns, A. C., Saxena, R., Vetter, C., Phillips, A. J. K., Lane, J. M., & Cain, S. W. (2021). Time spent in outdoor light is associated with mood, sleep, and circadian rhythm-related outcomes: A cross-sectional and longitudinal study in over 400,000 UK Biobank participants. J Affect Disord, 295, 347–352. 10.1016/j.jad.2021.08.056

8. Christensen, R. H. B. (2019). Ordinal - regression models for ordinal data. In R package version 2019.4-25. http://www.cran.r-project.org/package=ordinal/

9. CIE. (2018). CIE S 026/E:2018: CIE System for Metrology of Optical Radiation for ipRGC-Influenced Responses to Light. In. Vienna: CIE Central Bureau.

10. Danilenko, K. V., Stefani, O., Voronin, K. A., Mezhakova, M. S., Petrov, I. M., Borisenkov, M. F., Markov, A. A., & Gubin, D. G. (2022). Wearable Light-and-Motion Dataloggers for Sleep/Wake Research: A Review. Applied Sciences, 12(22). 10.3390/app122211794

11. Figueiro, M. G., Hamner, R., Bierman, A., & Rea, M. S. (2013). Comparisons of three practical field devices used to measure personal light exposures and activity levels. Light Res Technol, 45(4), 421–434. 10.1177/1477153512450453

12. Gaddy, J. R., Rollag, M. D., & Brainard, G. C. (1993). Pupil size regulation of threshold of light-induced melatonin suppression. J Clin Endocrinol Metab, 77(5), 1398–1401. 10.1210/jcem.77.5.8077340

13. Harley, S. K., & Sliney, D. H. (2018). Pupil Size in Outdoor Environments. Health Phys, 115(3), 354–359. 10.1097/HP.0000000000000887

14. Harris, P. A., Taylor, R., Minor, B. L., Elliott, V., Fernandez, M., O’Neal, L., McLeod, L., Delacqua, G., Delacqua, F., Kirby, J., Duda, S. N., & Consortium, R. E. (2019). The REDCap consortium: Building an international community of software platform partners. J Biomed Inform, 95, 103208. 10.1016/j.jbi.2019.103208

15. Harris, P. A., Taylor, R., Thielke, R., Payne, J., Gonzalez, N., & Conde, J. G. (2009). Research electronic data capture (REDCap)--a metadata-driven methodology and workflow process for providing translational research informatics support. J Biomed Inform, 42(2), 377–381. 10.1016/j.jbi.2008.08.010

16. Hartmeyer, S. L., Webler, F. S., & Andersen, M. (2022). Towards a framework for light-dosimetry studies: Methodological considerations. Lighting Research & Technology, 55(4-5), 377–399. 10.1177/14771535221103258

17. Higuchi, S., Ishibashi, K., Aritake, S., Enomoto, M., Hida, A., Tamura, M., Kozaki, T., Motohashi, Y., & Mishima, K. (2008). Inter-individual difference in pupil size correlates to suppression of melatonin by exposure to light. Neurosci Lett, 440(1), 23–26. 10.1016/j.neulet.2008.05.037

18. Hubalek, S., Zöschg, D., & Schierz, C. (2016). Ambulant recording of light for vision and non-visual biological effects. Lighting Research & Technology, 38(4), 314–321. 10.1177/1477153506070687

19. Lazar, R., Degen, J., Fiechter, A. S., Monticelli, A., & Spitschan, M. (2024). Regulation of pupil size in natural vision across the human lifespan. R Soc Open Sci, 11(6), 191613. 10.1098/rsos.191613

20. Lucas, R. J., Peirson, S. N., Berson, D. M., Brown, T. M., Cooper, H. M., Czeisler, C. A., Figueiro, M. G., Gamlin, P. D., Lockley, S. W., O’Hagan, J. B., Price, L. L., Provencio, I., Skene, D. J., & Brainard, G. C. (2014). Measuring and using light in the melanopsin age. Trends Neurosci, 37(1), 1–9. 10.1016/j.tins.2013.10.004

21. Marcos, S. (2025). Optical and Visual Diet in Myopia. Invest Ophthalmol Vis Sci, 66(7), 3. 10.1167/iovs.66.7.3

22. Okudaira, N., Kripke, D. F., & Webster, J. B. (1983). Naturalistic studies of human light exposure. Am J Physiol, 245(4), R613–615. 10.1152/ajpregu.1983.245.4.R613

23. Peeters, S. T., Smolders, K. C. H. J., & de Kort, Y. A. W. (2020). What you set is (not) what you get: How a light intervention in the field translates to personal light exposure. Building and Environment, 185. 10.1016/j.buildenv.2020.107288

24. Price, L. L. A., & Blattner, P. (2022). Circadian and visual photometry. Prog Brain Res, 273(1), 1–11. 10.1016/bs.pbr.2022.02.014

25. R Core Team. (2017). R: a language and environment for statistical computing. In R Foundation for Statistical Computing. https://www.R-project.org/

26. Sliney, D. H. (1995). UV radiation ocular exposure dosimetry. J Photochem Photobiol B, 31(1-2), 69–77. 10.1016/1011-1344(95)07171-5

27. Sliney, D. H. (1997). Ocular exposure to environmental light and ultraviolet--the impact of lid opening and sky conditions. Dev Ophthalmol, 27, 63–75. 10.1159/000425651

28. Sliney, D. H. (2002). How light reaches the eye and its components. Int J Toxicol, 21(6), 501–509. 10.1080/10915810290169927

29. Spitschan, M. (2019). Melanopsin contributions to non-visual and visual function. Curr Opin Behav Sci, 30, 67–72. 10.1016/j.cobeha.2019.06.004

30. Spitschan, M., Smolders, K., Vandendriessche, B., Bent, B., Bakker, J. P., Rodriguez-Chavez, I. R., & Vetter, C. (2022). Verification, analytical validation and clinical validation (V3) of wearable dosimeters and light loggers. Digit Health, 8, 20552076221144858. 10.1177/20552076221144858

31. Stampfli, J. R., Schrader, B., di Battista, C., Hafliger, R., Schalli, O., Wichmann, G., Zumbuhl, C., Blattner, P., Cajochen, C., Lazar, R., & Spitschan, M. (2023). The Light-Dosimeter: A new device to help advance research on the non-visual responses to light. Light Res Technol, 55(4-5), 474–486. 10.1177/14771535221147140

32. Thieden, E., Agren, M. S., & Wulf, H. C. (2000). The wrist is a reliable body site for personal dosimetry of ultraviolet radiation. Photodermatol Photoimmunol Photomed, 16(2), 57–61. 10.1034/j.1600-0781.2000.d01-4.x

33. Wen, L., Cheng, Q., Cao, Y., Li, X., Pan, L., Li, L., Zhu, H., Mogran, I., Lan, W., & Yang, Z. (2021). The Clouclip, a wearable device for measuring near-work and outdoor time: validation and comparison of objective measures with questionnaire estimates. Acta Ophthalmol, 99(7), e1222–e1235. 10.1111/aos.14785

34. Wen, L., Liu, H., Chen, Z., Xu, Q., Hu, Z., Lan, W., & Yang, Z. (2023). Effect of mount location on the quantification of light intensity in myopia study. BMJ Open Ophthalmol, 8(1). 10.1136/bmjophth-2023-001409

35. Windred, D. P., Burns, A. C., Lane, J. M., Olivier, P., Rutter, M. K., Saxena, R., Phillips, A. J. K., & Cain, S. W. (2024). Brighter nights and darker days predict higher mortality risk: A prospective analysis of personal light exposure in >88,000 individuals. Proc Natl Acad Sci U S A, 121(43), e2405924121. 10.1073/pnas.2405924121

36. Yoshimura, M., Kitamura, S., Eto, N., Hida, A., Katsunuma, R., Ayabe, N., Motomura, Y., Nishiwaki, Y., Negishi, K., Tsubota, K., & Mishima, K. (2020). Relationship between Indoor Daytime Light Exposure and Circadian Phase Response under Laboratory Free-Living Conditions. Biological Rhythm Research, 53(5), 765–785. 10.1080/09291016.2020.1782691

37. Zauner, J., Broszio, K., & Bieske, K. (2023). Influence of the Human Field of View on Visual and Non-Visual Quantities in Indoor Environments. Clocks Sleep, 5(3), 476–498. 10.3390/clockssleep5030032

38. Zielinska-Dabkowska, K. M., Schernhammer, E. S., Hanifin, J. P., & Brainard, G. C. (2023). Reducing nighttime light exposure in the urban environment to benefit human health and society. Science, 380(6650), 1130–1135. 10.1126/science.adg5277

